# HPV detection patterns in young women from the PAPCLEAR longitudinal study: implications for HPV screening policies

**DOI:** 10.1101/2023.09.30.23296382

**Authors:** Thomas Beneteau, Soraya Groc, Carmen Lia Murall, Vanina Boué, Baptiste Elie, Nicolas Tessandier, Claire Bernat, Marine Bonneau, Vincent Foulongne, Christelle Graf, Sophie Grasset, Massilva Rahmoun, Michel Segondy, Vincent Tribout, Jacques Reynes, Christian Selinger, Nathalie Boulle, Ignacio G. Bravo, Mircea T. Sofonea, Samuel Alizon

**Affiliations:** MIVEGEC, Univ Montpellier, CNRS, IRD, France; PCCEI, Univ Montpellier, Inserm, EFS, Montpellier, France; Department of Biological Sciences, Université de Montréal, Montréal, Canada; Center for Interdisciplinary Research in Biology (CIRB), Collége de France, CNRS, INSERM, Université PSL, Paris, France; Institut de Génomique Fonctionnelle, Université de Montpellier, CNRS, INSERM, Montpellier, France; Department of Obstetrics and Gynaecology, Centre Hospitalier Universitaire de Montpellier, Montpellier, France; Center for Free Information, Screening and Diagnosis (CeGIDD), Centre Hospitalier Universitaire de Montpellier, Montpellier, France; Department of Infectious and Tropical Diseases, Centre Hospitalier Universitaire de Montpellier, Montpellier, France; Swiss Tropical and Public Health Institute, Basel, Switzerland; CHU de Nîmes, Nîmes, France

**Keywords:** Papillomaviruses, Vaccination, Epidemiology, Public Health

## Abstract

**Objectives:** HPV infections are ubiquitous. For most infections, we lose track of the presence of the virus in host in less than three years after the start of infection. The mechanisms regulating the persistence of HPV infection are still partially understood. In this work, we focus on incident HPV detection in young women and we characterise the dynamics of these infections and evaluate the effect of genotype and host socio-economic factors on the duration of HPV detection and time between detection.

**Methods:** We investigated human papillomavirus (HPV) genotype detection patterns in 182 young women in Montpellier, France. We relied on SPF_10_-LiPA25 screening assay for the simultaneous detection of 25 HPV genotypes. We used survival analysis tools with frailty effects to investigate the contribution of viral and host factors to variations in the time of HPV detectability, time of first incident detection, and time before re-detection.

**Results:** Women of the PAPCLEAR cohort experienced numerous positive HPV events, including frequent redetection of the same genotype. We retrieve classical results that HR-genotypes are detected for longer duration than LR-genotypes. HR-genotypes were also more liekly to be detected than LR-genotypes during the follow-up. The number of lifetime sexual partner was strongly associated with increased risk of new positive detection while vaccination was related to a lower risk of displaying incident infections. Covariates related to socio-economic difficulties were associated with longer duration of HPV positivity.

**Conclusions:** Young women display numerous event of HPV detection, with frequent codetections of multiple genotypes at the same time and redetection of the same type after periods of no detection. These new detection are almost certainly the result of new acquisition from sexual partners, with little evidence of re-emergence of latent infections. A better characterisation of transient infections might help unveil doubts and misconception on HPV physiopathology, favouring adherence to preventive policies.

---

Human Papillomaviruses (HPV) are the most oncogenic viruses known to infect humans, accounting for more than 600,000 deaths worldwide each year [1]. They are also one of the most common sexually transmitted infections, with estimates suggesting that by 45yo, more than 80% of the people are or have already been infected by an HPV [2]. It is generally accepted that the initial HPV infection is acquired during the first sexual exposures, with the prevalence peaking after sexual debut, and that the risk of contracting a new HPV infection increases with the number of sexual partners [3]. HPV presence generally goes undetected within the first three years, an event generally referred as HPV clearance [4]. This clearance, however, may not necessarily imply true immune clearance. The interpretation of redetection events is delicate as they might originate from various sources: true new infection, transient deposition from a sexual partner, or detection of latent infection [5].

Most certainly, HPV detection is a combination of these different pathways. Deciphering the cause of new HPV detection is still a major challenge. Answering this question is crucial in the optimisation of future public health policies, to evaluate the effectiveness of catch-up vaccination or organise the age stratification of vaccination policies.

Longitudinal studies are valuable data both in terms of density and length of follow-up. Extracting full potential of such raw material is challenging and require the use of rigorous statistical tools. In this work, we used the PAPCLEAR cohort which of samples collected every 8 weeks in 132 young women, aged 18-25, for which we test the presence of 25 HPV genotypes using the SPF10-LiPA25 technique [6]. In particular, we evaluated viral genotypes and host factors involved with attention to frailty effects at the patient level and accounting for the censoring in the data to maximize the quality of the analysis.

## Materials and methods

### Study design and participants

The PAPCLEAR cohort has been detailed elsewhere [7]. In short, this monocentric longitudinal study included 189 women, who were between 18 and 25 years old at inclusion, lived in the area of Montpellier (France) and reported having at least one new sexual partner over the last 12 months. Women with a history of HPV-associated pathology were excluded from the study. Pregnant women or women who were planning a pregnancy within the first year of inclusion were also excluded from the study. A graphic summarising the inclusion protocol can be found in the Supplementary materials S4. A total of 150 women were followed longitudinally for up to 2 years between 2016 and 2020. The additional 39 participants were part of a cross-sectional analysis, that was prematurely suspended due to the pandemic. On-site visits of infected participants took place every 8 weeks with a gynaecologist or a midwife, who performed a cervical smear. Except for inclusion, participants were told to avoid sexual contact the day before the visit took place to avoid unwanted transient sexual deposition from the partner. At inclusion, the participants self-completed an extensive questionnaire related to demographic, socioeconomic, and behavioural risk factors. For the next visits, the participants also filled in follow-up questionnaires to notify changes in their habits. All participants provided written informed consent.

### Genotyping

We first tested for the presence of alpha papillomaviruses in the cervical smears using the DEIA assay [8]. DEIA-positive samples were genotyped using the LiPA25 assay, which was chosen for its sensitivity and can detect up to 25 different HPV genotypes [6]. Among these, we refer to high-risk (HR) genotypes for HPV16, 18, 31, 33, 35, 39, 45, 51, 52, 56, 58, 59, 66, 68 [9] and to low-risk (LR) for the remaining 12 (HPV6, 11, 34, 40, 42, 43, 44, 53, 54, 70, 74). If the LiPA25 test was negative, the genotype was determined using the PGMY PCR amplification [10] and Sanger sequencing of the PCR product. If the sequencing did not yield a clear result, samples were labelled as ‘non-typable’.

#### Statistical analyses

All statistical analyses were conducted using R 4.2.2 with additional packages listed in Supplementary materials A6.

We excluded all the women with less than three visits, meaning all the cross-sectional group and 18 participants from the longitudinal group (1 participant quit the study, 4 were seen once, and 13 were seen only twice). All analyses were genotype-specific, with the unit of observation being the HPV genotype. Therefore, each participant could contribute to multiple observations. Following earlier studies, we assumed the dynamics of each genotype to be independent at the participant level and between participants [11]. If not specified, the results were pooled across all genotypes.

As used in previous works [12], we defined an HPV genotype as ‘prevalent’ if detected at inclusion. We also defined a genotype as ‘incident’ if detected at posterior visits but not at inclusion. Patterns of positive detection separated by only one negative visit, sometimes referred to as ‘intermittent detection’ [12], are handled differently between studies and there does not appear to be consensus on the way to deal with such data. In the main analysis, we considered the two episodes as separated but we also conducted analysis with mergedintermittent patterns (Supplementary materials A5).

Model selection was performed using the corrected Akaike Information Criterion (AICc) as a metric for the penalised goodness of fit [13] We evaluated the goodness of fit for all sub-models of the maximum model (i.e. with all the covariates) and estimated the hazard ratios of the Cox regression using a full averaging procedure on the best models. Thorough details are available in the Supplementary materials A4. To test for differences between the two populations, we used Fisher’s exact test for qualitative variables and Wilcoxon-Mann-Whitney’s test for quantitative variables. We displayed the raw *p*-values in the results Tables and Figures. In the following, a p-value *<* 0.05 is considered significant.

### Survival analyses

For each genotype and each participant, we analysed the duration of HPV detectability, the duration between HPV-positive episodes, and the time to incident HPV infection. Survival functions for these quantities were computed using the non-parametric Nelson-Aalen estimator [14, 15]. For the time until the first incident detection, we fitted a Weibull distribution to the survival function to predict the cumulative risk of incident detection at 5 years since inclusion (see the Supplementary materials A7 for more details).

For a given episode, we defined the time of HPV detectability as the duration between the midpoint at the start of the episode and the midpoint at the end of the episode. We included all incident episodes, even the shortest episodes that were only detected during one visit, elsewhere referred to ‘transient’ infections, but hereafter called ‘singletons’, and the right-censored observations. The latter corresponds to patients who tested positive for HPV at their last scheduled visit. We also included prevalent episodes whose start is unknown and for which the duration of HPV detectability is right-censored. Similarly, the time between consecutive positive episodes was computed as the duration between the midpoint at the end of the expired episodes and the midpoint at the start of the new episodes. The time until the first incident infection here corresponds to the time from inclusion to the midpoint at the start of the first incident detection. Extensive information can be found in Supplementary Methods A3.

We checked for differences in HPV detectability and time to first incident detection between HR-genotypes and LR-genotypes using log-rank tests [16, 17]. To evaluate the effect of non-viral variables, we used Cox proportional hazards models [18]. We stratified the Cox regression with different baseline hazard functions for genotypes not detected, first detection, first redetection, and second redetection. We assumed no interaction between the strata. For all Cox regression analyses, we checked the validity of the proportional hazards (PH) assumption using Schoenfeld’s residuals [19]. The covariates included in the analysis are the number of lifetime sexual partners (LTSP), the BMI at baseline, the self-declared ethnic origin (Caucasian vs. non-caucasian or mixed-origin), the HPV vaccination status, the sexual affinity, the use of condom or contraceptive pills, an indicator of financial difficulties (participant had to decline medical care because of financial reasons), the number of years between inclusion and first menstruation, the number of years between inclusion and first sexual intercourse and the smoking stats (past, current or never). The numbers for each category can be found in Table 1.

**Table 1:** Baseline description of the PAPCLEAR cohort for participants included in the analysis (. *>* 2 **visits).** Except for the vaccine used not included in the analysis, the first level display for all categorical variables is the reference level used in the Cox analysis. Missing observations were removed from the analysis.

| Covariates | States/mean [IQR] | # women | % women |
| --- | --- | --- | --- |
| Age at inclusion | 21.27 [20 - 24] | 132 | 100 |
| Age of first menstruation | 12.85 [12 - 14] | 131 | 99.24 |
| Years between 1st menstruation and inclusion | 8.42 [7 - 10] | 131 | 99.24 |
| Age of first sexual intercourse | 16.39 [15 - 17] | 132 | 100 |
| Years between 1st intercourse and inclusion | 4.89 [3 - 7] | 132 | 100 |
| Vaccination | Unvaccinated | 66 | 50.00 |
|  | Vaccinated | 66 | 50.00 |
| Vaccine used | Cervarix | 6 | 9.09 |
|  | Gardasil | 56 | 84.18 |
|  | Non-specified | 4 | 6.06 |
| Self-declared ethnicity | Mixed or other | 27 | 20.45 |
|  | Caucasian | 105 | 79.55 |
| Self-declared sexual affinity | Bi-/Homosexual | 12 | 9.09 |
|  | Heterosexual | 120 | 90.90 |
| Smoking | Never | 61 | 46.21 |
|  | Past | 19 | 14.39 |
|  | Current | 52 | 39.39 |
| Contraceptive pills <sup>†</sup> | Not using | 64 | 48.48 |
|  | User | 68 | 51.51 |
| Male condoms | Not using | 56 | 42.42 |
|  | User | 76 | 57.57 |
| Number of lifetime sexual partner | 1;2 | 20 | 15.15 |
|  | 3;10 | 63 | 47.72 |
|  | 11+ | 48 | 36.36 |
|  | missing | 1 | 0.76 |
| Financial difficulties <sup>*</sup> | No experience | 117 | 88.64 |
|  | Experienced in the last 12mo | 15 | 11.36 |

<sup>†</sup> emergency pills not included
<sup>\*</sup> defined as a participant who declined medical care because of financial reasons

We considered here that the unit of observation was the HPV genotype at the patient level. Thus as one participant would experience codetection of multiple genotypes, this could induce some correlations between the observations. To account for correlations between observations of the same cluster (i.e. a participant), we add shared frailty effects at the patient level in the Cox regression [20]. We tested for the relevance of adding the frailty at the patient level using a likelihood ratio test with one degree of freedom between the two models (with and without the frailty effect).

## Results

### Descriptive analysis

The 150 participants of the PAPCLEAR cohort came for 6 visits on average (Poisson 95% CI: 5.63 - 6.43). For the 132 women included in the analysis, the follow-up duration was on average 311 days (IQR: 182 - 431), this accounted for a total of 1543 months of follow-up. The PAPCLEAR participants included in the analysis were on average aged 21.3yo (IQR: 20 - 23) at inclusion and around half of them (62/132) were vaccinated against HPV (56 with Gardasil and 6 with Cervarix). Baseline characteristics for the 132 women included can be found in Table 1.

In about 74% of the women included in the analysis (98/132), we detected at least one episode during their follow-up and 47% (62/132) experienced codetections (i.e. the simultaneous detections by more than one genotype). Overall, we detected 342 distinct episodes, 186 incident (including 137 first detections, 44 redetections and 5 second redetections) and 156 prevalent, including 211 from HR types and 114 from LR types. For 17 episodes, we could not determine the genotype detected. The three most frequent detected types in descending order were HPV51, HPV53, and HPV66, in agreement with previous results on the PAPCLEAR cohort [7]. A total of 83 (62.9%) participants were positively detected for at least one HPV genotype.

An average of 2.6 (Poisson 95% CI: 2.32-2.88) episodes per woman were detected during the whole follow-up, which yielded an average attack rate of 2.99 HPV episodes detected per person-year.

### Detection of first incident HPV episode

Overall, we detected 137 first incident detection for all detectable genotypes and all participants. At one year, we estimated a 4.80% (log-log 95% CI: 3.99 - 5.77) cumulative proportion of first incident HPV detection pooled across all genotypes. After two years of follow-up, the proportion of first incident infection detection increased to 7.26% (log-log 95% CI: 5.84 - 9.02). To assess the variation after 5 years, we used the Weibull fit and predicted the proportion to reach 16.56% (95% CI: 7.37 - 29.73). Information regarding the Weibull fit can be found in the Supplementary materials A7.

We assessed the rate of incident detection by oncogenic risk. We found that HR-HPV were more likely to be detected than LR-HPV over the whole follow-up (log-rank p-value *<* 0.01). We, however, lacked statistical power to verify that the difference in survival functions between HR genotypes and LR genotypes was consistent for redetection or second redetection.

### Risk factors for the time between consecutive detection

In addition to the 137 observed first incident detection (3271 right-censored), we detected 156 prevalent episodes, 44 observed redetection (213 right-censored) and 5 second redetection (36 right-censored). Among the redetection, 33 consecutive episodes were only separated by one negative visit, pattern else-where referred to as ‘intermittent’ detection[12]. Compared to participants reporting 1 or 2 LTSP, women who reported 3-10 LTSP had increased risk of experiencing new detection(hazard ratio: 2.40; 95% CI:1.07 - 5.39), first incident or redetection. We observed a similar trend for participants reporting more than 10 LTSP, compared to the reference of 1 or 2 LTSP. However, we lacked statistical power to report significant association (hazard ratio: 2.15; 95% CI:0.94 - 4.93). Merging intermittent patterns (Figure S2) yielded similar results, this time with a significant association for the group reporting more than 10 LTSP. We also found that vaccinated participants were less likely to display new incident detection or redetection compared to unvaccinated participants (hazard ratio: 0.64; 95% CI: 0.43-0.96). Thorough results of the Cox analysis for the time between consecutive episodes are displayed in Figure 1.

**Figure 1:**
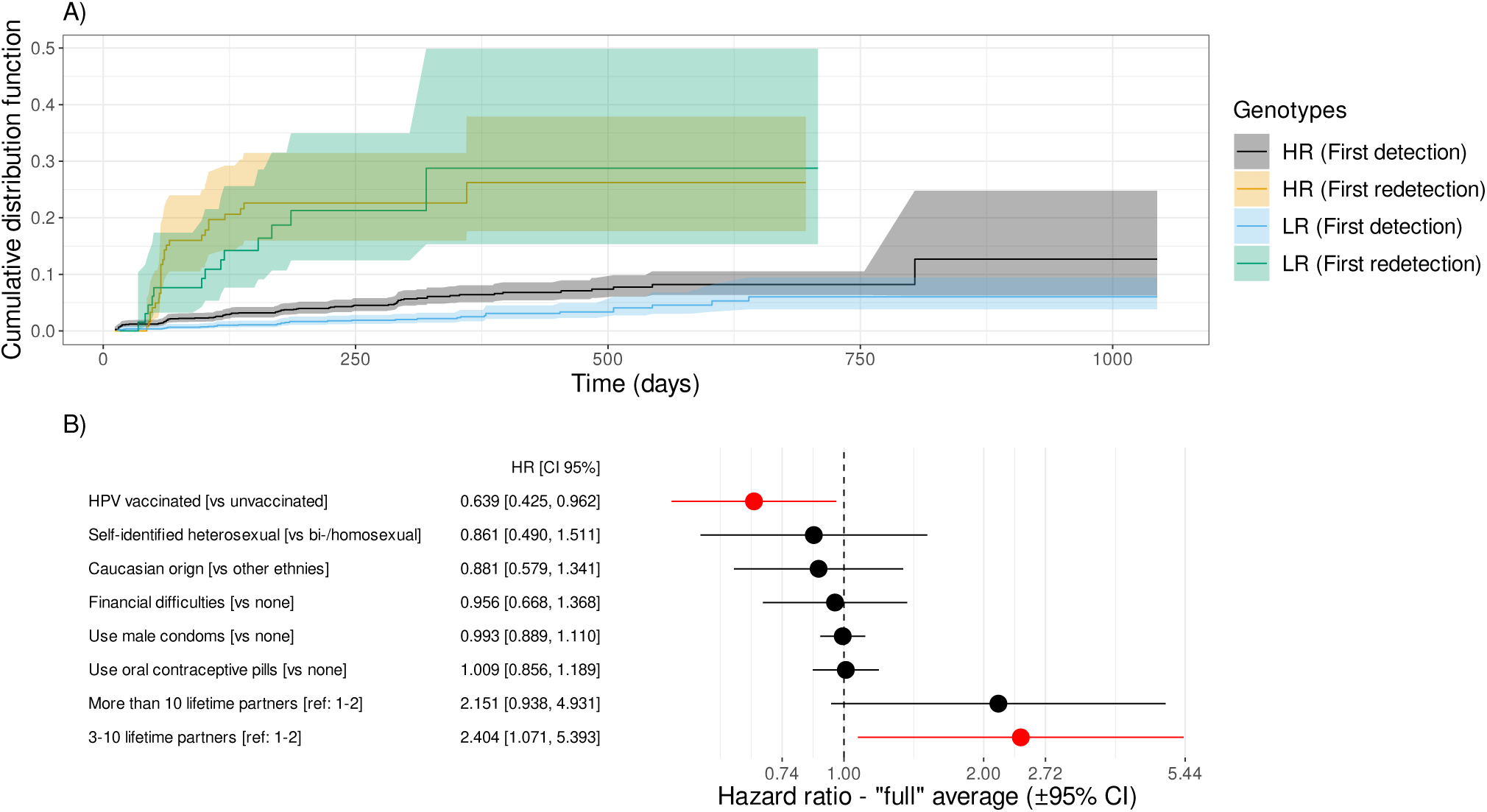
Cumulative distribution functions for the time to first incident detection and time to first genotype redetection stratified by HR/LR genotypes and effects of host covariates on these estimates. A) Cumulative distribution function (CDF) of the time to fist incident HPV detection since inclusion and the time to the first redetection, stratified by HPV genotype status (HR and LR). B) Hazard ratio for the best models selected by Cox regression with frailty at the patient level. Significant covariates are in red and hazard ratios greater than 1 indicate the covariate is associated with an increased risk of detection, hence lower duration between consecutive episodes.

### Loss of HPV detection

On the total of 342 detected episodes, 156 were prevalent HPV detection and for 40 episodes we did not observe the loss of detectability. For 17 episodes, the participants entered positive to a genotype and left the follow-up still positive for that same genotype, without any negative visit in-between. The majority of the episodes were positive for only one visit (198/342; 57.9%), but a significant proportion of them are censored observations (86/198; 43.4%) and thus potentially just a glimpse of a longer event. We estimated a median period of HPV detectability of 113 days (log-log 95% CI: 92.5 - 124). Our results suggest that around 23.5% (log-log 95% CI: 16.5 - 31.4) of the HPV episodes were still detectable after 700 day of follow-up. We found that HR-HPV types were detected significantly longer than LR-HPV infections (log-rank p-value *<* 0.05), the survival functions are displayed in Figure 2. The median duration of detectability was 130 days (log -log 95% CI 106 - 186) for HR genotypes and 96 days (log-log 95% CI 67.5 - 113) for LR genotypes.

**Figure 2:**
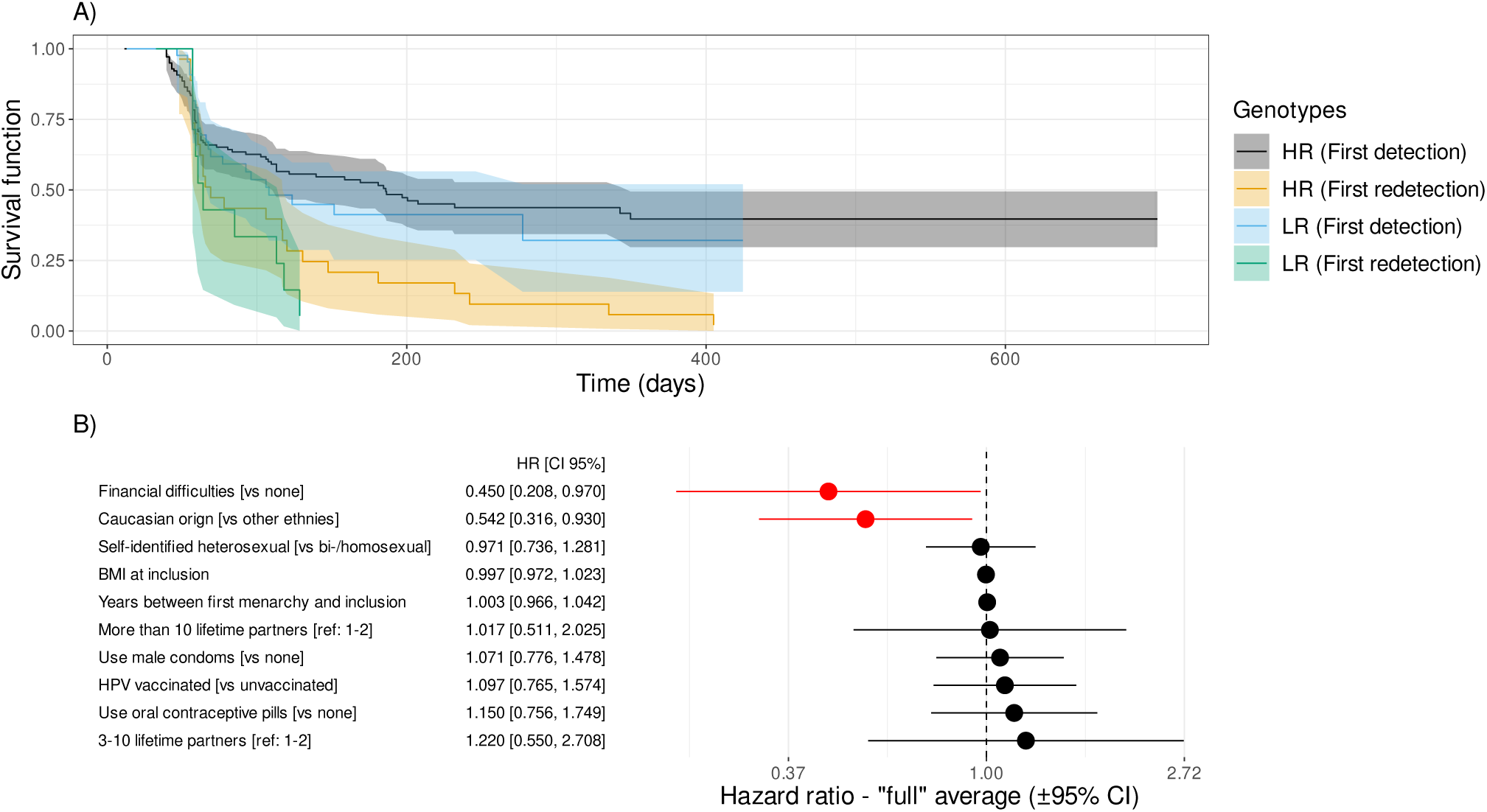
Survival function for the time of HPV detectability stratified by HR/LR genotypes and effects of host covariates on this estimate. A) Survival functions stratified by the genotype (HR/LR) for the time to loss of HPV detection. B) Hazard ratios for the host factors. Significant factors are in red and a hazard ratio lower than one indicates that the trait is associated with a decreased rate of loss of HPV DNA detection, hence longer survival functions. The reference level is indicated in the bracket for the qualitative variables (see Methods for details).

Finally, we investigated the effect of key host factors (listed in Table 1) on the duration of HPV detectability using LiPA assays. Our analysis showed that HPV was detected for a significantly longer duration in participants who experienced financial difficulties (defined as a participant who declined medical care because of financial reasons) in the last 12mo before inclusion compared to participants who did not experience it (Hazard ratio: 0.45; 95% CI: 0.21 - 0.97). Infections were also detected longer in participants who identified themselves solely as ‘Caucasians’ (Hazard ratio: 0.54; 95% CI: 0.32 - 0.96) compared to participants who identified themselves as non-Caucasian or mixed-origin (Figure 2). These results were similar when merging intermittent patterns (Figure S3).

## Discussion

In this work, we analysed the HPV detection patterns of 132 young women from the PAPCLEAR cohort. We estimated a cumulative probability of first incident HPV detection since baseline, pooled across all genotypes, of 4.80% (log-log 95% CI: 3.99 - 5.77) at one year and predicted it to reach 16.56% (95% CI: 7.37 - 29.73) at five year. While we used convenient denominations such as ‘first incident detection’ or ‘first redetection’ following previous works, these expressions surely not bare the same relevance in terms of natural history. It is very unlikely for most of the investigated participants that a first detection during the follow-up corresponds to a true first exposure, only 11.36% participants declared having had their first sexual partner in the last 12 months. Besides, these definitions are dependent of the sampling frequency, as the probability of documenting a short period between redetections increases with the sampling rate. Thus, it is not straightforward to compare our results with other studies. Our results for first detection are very similar to results for first redection but quite discordant with estimates for first incident detection from other studies [12]. Those differences might first come from the difference in the cohort design, as the sampling rate for the Ludwig-McGill cohort was about every 6 months. Besides, the two populations were quite different. Participants here are all between 18-25 years old at inclusion. In the latter, there is a wider age diversity among the women, with around 80% of the women being older than 25 years old. Sexual activity is negatively correlated with age after sexual debut, thus our population might be more exposed to HPV due to increased sexual activity [21]. Additionally, the Ludwig-McGill cohort was sampled from low-income families from Brazil, while we included women without income criteria.

We found that the participants experienced numerous detected episodes (2.99 per women-year), most of them being positively detected for only one visit (198/374 singletons, 86/198 censored). In a quarter of the participants included in the analysis (34/132), no alphapapillomaviruses was detected, while for 18 women we detected more than 5 episodes during the follow-up. Redetection, were not uncommon as in about a third of women displaying at least one HPV positive visit we detected redetections of the same genotype. The frequency of codetections is similarly high (47%) and is consistent with previous studies [22].

To date, except for some specific populations (e.g. abstinent women [23]), it is quite difficult to settle on the origin of new HPV detection in humans. In our cohort, the number of lifetime partners was negatively associated with the duration between episodes. We found that in participants reporting a number of lifetime partners higher than 3, the time between episodes was significantly shorter than for women with 1 or 2 reported LTSP at inclusion, with little difference between participants reporting 3 to 10 LTSP and those reporting more than 11 LTSP. The number of LTSP was not associated with the number of years between first intercourse and inclusion. Besides, women reporting a higher number of LTSP at inclusion were also more likely to report intercourse with new partner during the follow-up. Taken together, our results suggest that new detection and redetection observed here are more likely to be new acquisition than re-emergence of latent infection. Additionally, we recall that only women who declared having sex with a new partner in the last 12 months were included in the cohort [7]. It was noted elsewhere that in the setting of new sexual partners, true incident infection was the preferential explanation [24, 25]. Compared to unvaccinated participants were less at risk of displaying incident infections. These results are consistent with observations in a dozen country [26].

We found that HR-HPV types were more likely to be detected for a longer period than LR-HPV types, which corroborates earlier studies [27]. Conversely, we found that the time between HPV positive events was shorter for HR-HPV types compared to LR-HPV types, consistent with other work [22].

The mean duration of HPV detectability was globally lower than previous studies [12, 22, 23, 27]. This can be partially explained by the difference in sampling rates with compared to studies (8 weeks compared to 6 months) and the inclusion of all positive episodes, including the singletons, sometimes excluded from analysis [23]. Participants that experienced financial difficulties (defined as a participant who declined medical care because of financial reasons) in past 12 months prior to inclusion displayed longer periods of HPV detectability compared to participants that did not declared suffering from financial struggles. People in situations of poverty generally tend to live in areas with low medical coverage, thus also limiting their access to medical care [28]. While there was no significant difference in vaccination uptake between participants who faced financial difficulties and those who did not, taken together, our results suggest that people with financial difficulties might be less prone to seek medical guidance, especially since specialists are not fully reimbursed in France, thus putting their more at risk of genital infections and complications. Our results also suggest that self-declared Caucasian participants experienced longer periods of HPV detection. However, we lacked information to assess if that trend originated from genetic origin or socio-demographic/behavioural differences between the two groups. Besides, our population is relatively limited in number (132 women, 27 mixed origin or non-Caucasian origin, 105 Caucasian origin) and restricted to a specific region in France. It does not reflect the French population diversity, and thus might just be the results of selection bias.

Clarifying the dynamic of HPV infection, especially regarding the distinction between re-detection and new acquisition is decisive to inform public health policies. Efficient screening policies and prevention have been implemented to limit progression towards cancer with good compliance to these measures [29]. While most HPV infections are generally benign, testing positive during HPV screening can cause psychological stress and anxiety [30], especially if self-sampling becomes widespread [31].

We believe a better characterisation of HPV infections, especially regarding the link between infection status and detection data, will help unveil doubts and misconception on HPV physiopathology, favouring adherence to preventive measures [32].

## Data Availability

The raw data and R scripts used will be deposited on the Zenodo server upon publication.

## Acknowledgements

The authors thank all the participants of the PAPCLEAR study and the clinical staff and nurses for their help.

## Supplementary Materials

### A1 Ethics

The PAPCLEAR trial was promoted by the Centre Hospitalier Universitaire de Montpellier and approved by the *Comité de Protection des Personnes* (CPP) *Sud Méditerrańee* I on 11 May 2016 (CPP number 16 42, reference number ID RCB 2016A00712-49); by the *Comité Consultatif sur le Traitement de l’Information en matìere de Recherche dans le domaine de la Santé* on 12 July 2016 (reference number 16.504); and by the *Commission Nationale Informatique et Libertés*on 16 December 2016 (reference number MMS/ABD/AR1612278, decision number DR-2016488). This trial was authorised by the *Agence Nationale de Śecurité du Médicament et des Produits de Santé*on 20 July 2016 (reference 20160072000007). The ClinicalTrials.gov identifier is NCT02946346. All participants provided written informed consent.

### A2 Protocol of PAPCLEAR study

**Figure S1:**
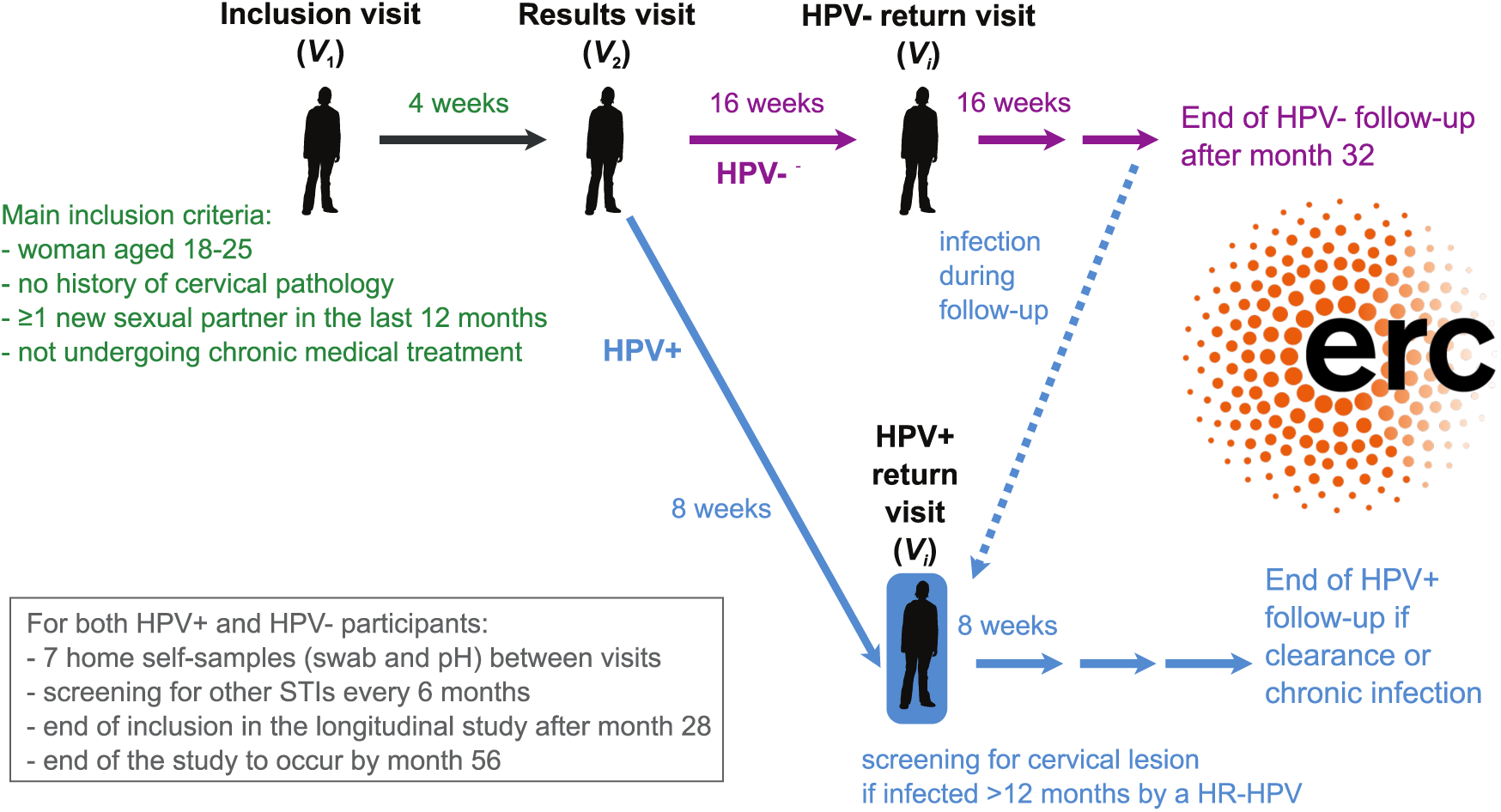
General structure of the PAPCLEAR study. [7].

### A3 Defining events and durations

Results from the DEIA and LiPA25 assays yielded dated binary vectors. For each infectious event, we only know the intervals during which the infection started and ended, which means the data is ‘doubly interval censored’ and usually cumbersome to analyse [33]. To simplify the problem, we computed duration using the conventional midpoint methodology. For this, we defined the start of an infection as the midpoint between the last negative test before and the first positive test of the infection. Likewise, we defined the end of an infection as the midpoint between the last positive test of the infection and the first negative test after the infection). For incomplete data, we assume the start to be at inclusion for left-censored observation and we assumed the end to be at last visit for right-censored observation. The bias associated with this simpler method is expected to be limited since our sampling scheme is regular and short-spaced [34].

To study the time to HPV infection clearance, we defined as an ‘event’ or ‘episode’ a series of at least one positive LiPA25 detection for a given HPV type and a given participant. During the follow-up, we often detected several events per participant (sometimes even by the same genotype). We assumed that two consecutive episodes were independent even if only separated by one negative visit. Such patterns, also called intermittent [12], are sometimes merged to form a longer episode instead of two separate entities [23]. We evaluated the changes in the estimates using this methodology below XXXX.

To estimate the time of HPV detectability, i.e. the time of positive HPV detection, we computed the duration between the midpoint at the start of an infection and the midpoint at the end of the infection. If one or both of the endpoints were censored, we assumed the duration to be right-censored. We assumed the events to be independent [11] and, therefore, defined the time between episodes to be independent events. For the time to first incident infection, we excluded prevalent infection and computed the time from inclusion to the midpoint at start of first incident detection for a genotype and a participant. If the genotype is not detected during follow-up, we used a right-censored observation whose duration equals the time of follow-up of the participant. When analysing the time between positive episodes, we included all events and computed the time as the duration between the midpoint at start of expired episodes and the midpoint at start of the new episodes. There is in general, a lower number of data of redetection than expected because some participant were still positive for a genotype at end of follow-up, thus preventing us from computing a time of redetection. In both cases, the cumulative distribution functions (CDF) or survival functions were computed using the Nelson-Aalen estimator of the cumulative hazard rate function [14, 15, 35].

### A4 Model comparison

We compared the models using the corrected Akaike Information Criterion (AICc) as a metric for the penalised goodness of fit [13] . Briefly, we first generated the maximum model with all the variables chosen for the Cox regression and then performed the model selection by subsetting all possible combinations from the maximum model and evaluating their respective AICc. We kept the models with an AICc smaller than the minimum AICc+2, following standard practice [36]. We then averaged the coefficients of the remaining models using a full averaging procedure to avoid artificial departure from 0. This was necessary because we averaged on all the selected models, not just on the ones with the variable whose coefficient was computed [37]. Finally, we computed the hazard ratio by taking the exponential of these averaged coefficients.

### A5 Merging intermittent patterns

Following previous notations, intermittent patterns corresponds to successive positive HPV detection episodes separated only by one negative visit. Merging intermittent patterns modifies the data used for analysis, diminishing the number of events, making them last longer in average. In total, we detect 33 intermittent patterns. Merging the patterns decreased the number of positive detectable events by the same amount. However, it did not change much the results of the Cox regression. While the degree of significance varies between the two datasets, the same trends were observed between the two cases.

**Figure S2:**
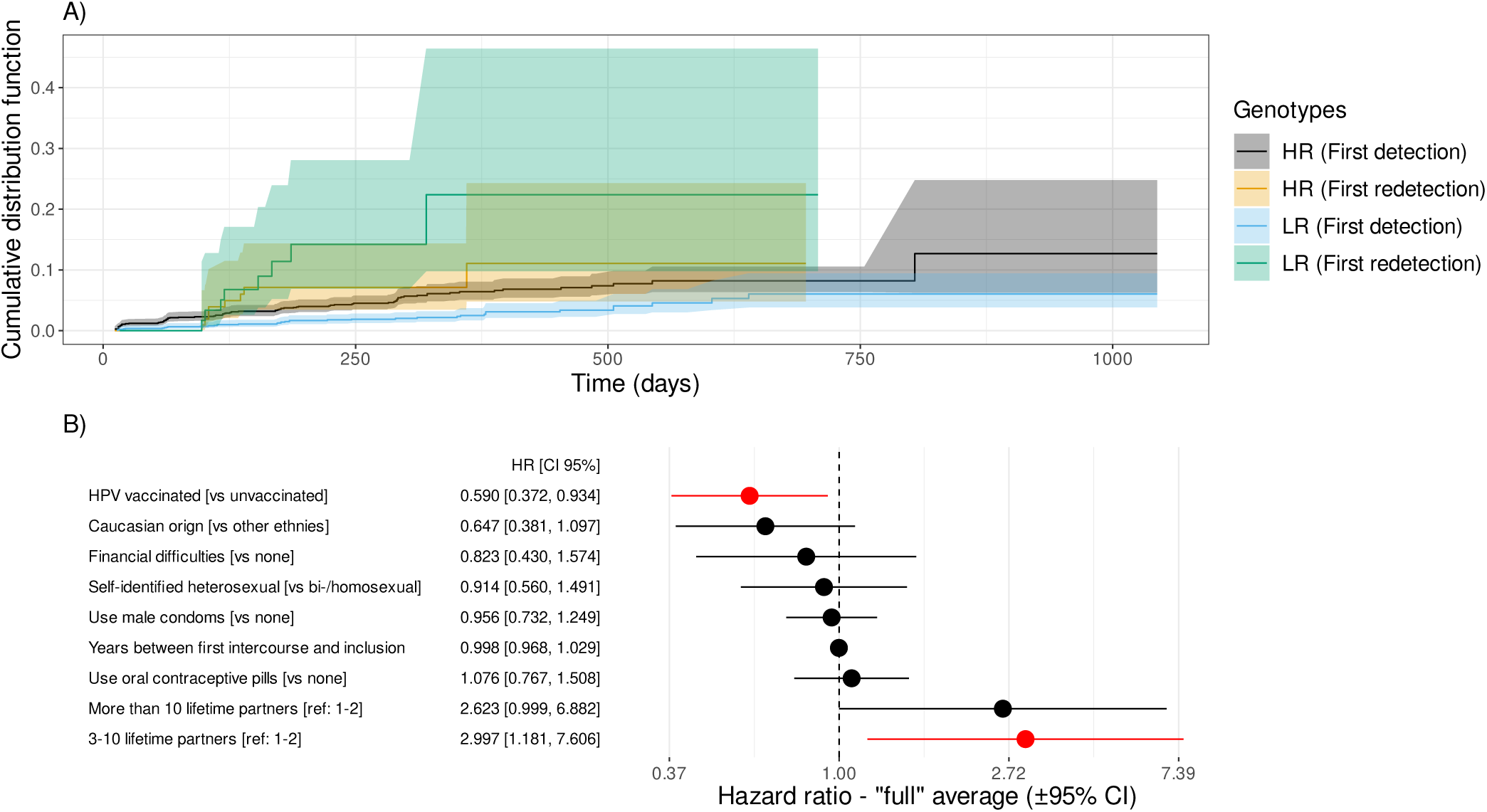
Cumulative distribution functions for the time to first incident detection and time to first genotype redetection stratified by HR/LR genotypes and effects of host covariates on these estimates for merged intermittent patterns. A) Cumulative distribution function (CDF) of the time to fist incident HPV detection since inclusion and the time to the first redetection, stratified by HPV genotype status (HR and LR). B) Hazard ratio for the best models selected by Cox regression with frailty at the patient level. Significant covariates are in red and hazard ratios greater than 1 indicate the covariate is associated with a increased risk of detection, hence lower duration between consecutive episodes.

**Figure S3:**
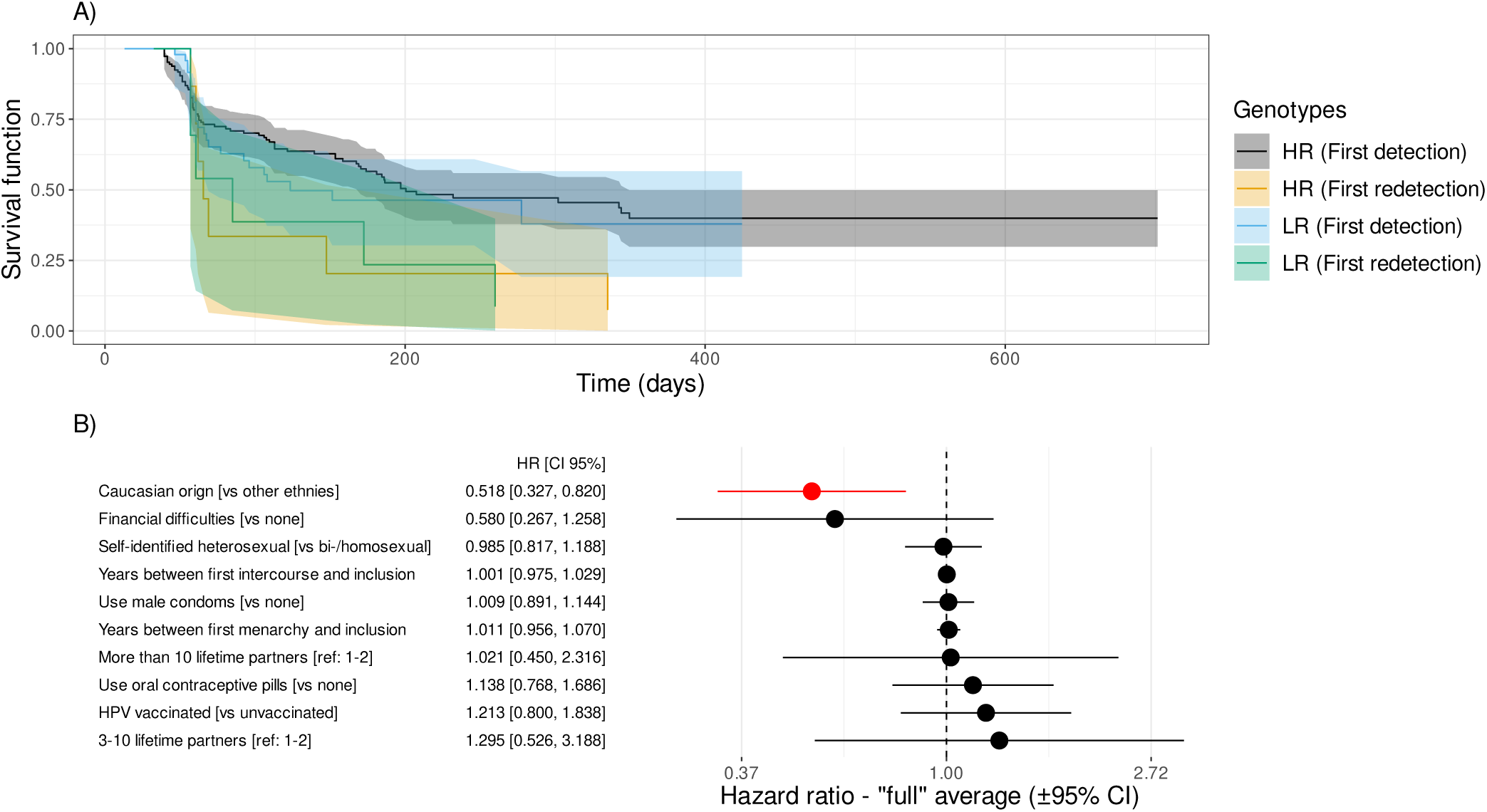
Survival function for the time of HPV detectability stratified by HR/LR genotypes and effects of host covariates on this estimate for merged intermittent patterns. A) Survival functions stratified by the genotype (HR/LR) for the time to loss of HPV detection. B) Hazard ratios for the host factors. Significant factors are in red and a hazard ratio lower than one indicates that the trait is associated with an decreased rate of loss of HPV DNA detection, hence longer surival functions. The reference level is indicated in the bracket for the qualitative variables (see Methods for details).

### A6 R packages

– survival: non-parametric and parametric estimators of the survival function and Cox regression [38]; version 3.5-3.
– MuMIn: model selection and model averaging [39]; version 1.46.0.
– coxme: adding frailty effects to the hazard function as a centred Gaussian distribution [40]; version 2.2-17.

### A7 Graphical Weibull fit

Let *λ >* 0 be the scale parameter and *k >* 0 be the shape parameter, for all *t ∈ R*^+^, we can define the survival function of the Weibull distribution as:

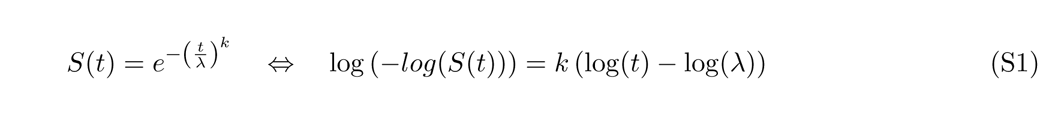

Thus using the *log*(*−log*(*·*)) transformation of the survival function, estimated using non-parametric estimators like Nelson-Aalen or Kaplan-Meier, and plotting it versus the natural logarithm of the event times, we can assess if a Weibull distribution is an appropriate model to describe the data by evaluating the goodness of the fit as a linear model [41]. Clearly, for the time to first incident detection pooled across all genotypes, and for both HR genotypes and LR genotypes grouping, the Weibull was relevent (panel A in Figure S4). However, for the time to loss of HPV detection, we see a clear non-linear trend between the *log*(*−log*(*·*)) transformation of the survival function and the log(time) (panel B in Figure S4), thus discouraging us for trying to fit a Weibull distributions to this data. The parameter estimates of the Weibull distribution for the time to first incident detection are displayed in Table S1.

**Figure S4:**
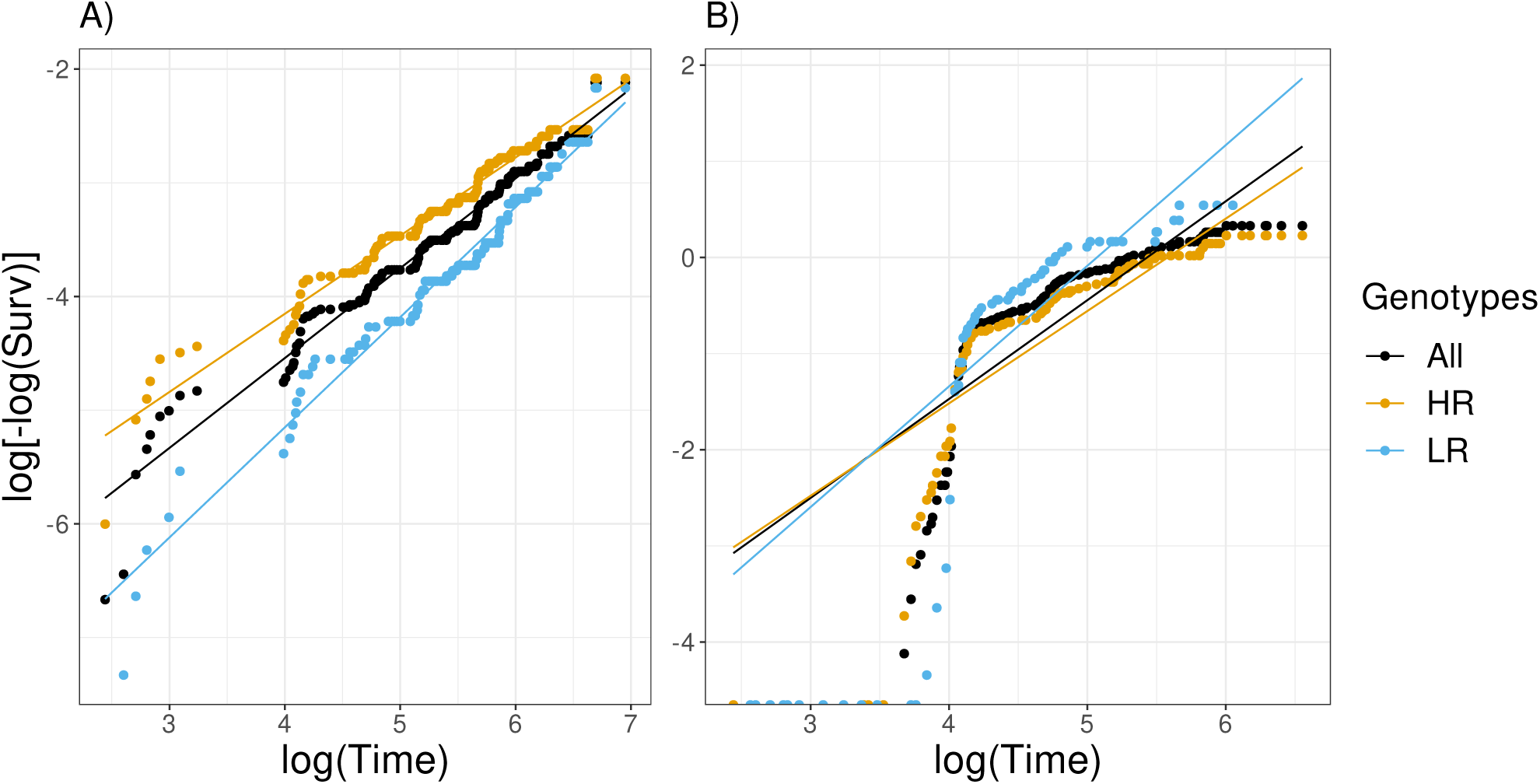
Graphical assessment for the goodness of Weibull fit. On panel A) we displayed the *log*(*−log*(*·*)) transformation of the survival function for the time to first incident detection versus the *log*(Time) and on panel B) the same transformation for the survival functions of the time to loss of HPV detection. For panel A) the linear fit is acceptable while for panel B) there is a clear non-linear trend.

**Table S1:** Estimates for the Weibull parameters (shape and scale) for the time to first incident detection.

| | scale ($\lambda$) | shape ($k$) |
| --- | --- | --- |
| All genotypes | $1.43 [0.79; 2.58] \times 10^4$ | $0.832 [0.713; 0.970]$ |
| HR genotypes | $1.12 [0.63; 2.01] \times 10^4$ | $0.832 [0.714; 0.971]$ |
| LR genotypes | $1.93 [1.27; 2.94] \times 10^4$ | $0.832 [0.714; 0.971]$ |

